# Characterizing subgroup performance of probabilistic phenotype algorithms within older adults: A case study for dementia, mild cognitive impairment, and Alzheimer’s and Parkinson’s diseases

**DOI:** 10.1101/2022.09.20.22280172

**Authors:** Juan M. Banda, Nigam H. Shah, Vyjeyanthi S. Periyakoil

## Abstract

**Objective:** Biases within probabilistic electronic phenotyping algorithms are largely unexplored. In this work, we characterize differences in sub-group performance of phenotyping algorithms for Alzheimer’s Disease and Related Dementias (ADRD) in older adults.

**Materials and methods:** We created an experimental framework to characterize the performance of probabilistic phenotyping algorithms under different racial distributions allowing us to identify which algorithms may have differential performance, by how much, and under what conditions. We relied on rule-based phenotype definitions as reference to evaluate probabilistic phenotype algorithms created using the Automated PHenotype Routine for Observational Definition, Identification, Training and Evaluation (APHRODITE) framework.

**Results:** We demonstrate that some algorithms have performance variations anywhere from 3 to 30% for different populations, even when not using race as an input variable. We show that while performance differences in subgroups are not present for all phenotypes, they do affect some phenotypes and groups more disproportionately than others.

**Discussion:** Our analysis establishes the need for a robust evaluation framework for subgroup differences. The underlying patient populations for the algorithms showing subgroup performance differences have great variance between model features when compared to the phenotypes with little to no differences.

**Conclusion:** We have created a framework to identify systematic differences in the performance of probabilistic phenotyping algorithms specifically in the context of ADRD as a use case. Differences in subgroup performance of probabilistic phenotyping algorithms are not widespread nor do they occur consistently. This highlights the great need for careful ongoing monitoring to evaluate, measure, and try to mitigate such differences.

## INTRODUCTION

The widespread adoption of machine learning (ML) algorithms for risk-stratification has unearthed plenty of cases of racial/ethnic biases within algorithms— from x-ray images to electronic health records (EHR) and clinical notes. [1–5]. When built without careful weightage, calibration, and bias-proofing, ML algorithms can give wrong recommendations, thereby worsening health disparities faced by communities of color. Medical researchers in fields like dermatology [6], pharmacovigilance [7], and clinical-decision support [8], to name a few, have started to examine biases inherently embedded within ML algorithms via the features used, quality of datasets, types of machine learning algorithms, and design decisions. Until the beginning of 2022, there have been over 600 published papers in PubMed, that address the evaluation and mitigation of racial bias in clinical ML models [9], with some pieces providing very insightful ideas (e.g. dividing bias in statistical and social) [10], listing challenges (e.g. adaptive learning, clinical implementation, and evaluating outcomes) [11], as well as strategies (e.g. reporting clarity, using de-noising strategies, explainability, among others) [12] on how to think about bias, where can it be present [13], and how it can be mitigated. On the implementation/deployment side, researchers have proposed how to introduce/represent these models to end-users [14] and some best practices within the field [15–18].

In the broader machine learning community, Kleinberg et al. [19] showed that a probabilistic classification to be ‘fair’ to different groups should satisfy three inherent conditions: 1) Calibration within groups, 2) Balance for the negative class, and 3) Balance for the positive class. However, these conditions cannot be satisfied all at once, which has led to the development of numerous other ‘fairness measures’ [20–22] that overlap and create confusion [23]. While most of these metrics apply to algorithms directly, they have not been analyzed in the context of medicine [24–26] until late 2019, with mixed and at times contradictory findings. When applied to medicine,other factors need to be considered, such as the clinical utility and benefit of the model [27].

Rule-based phenotyping has been the de facto method for identifying cohorts of patients belonging to any given condition/phenotype [28]. This method requires clinicians to agree on a set of clinical elements organized in logical rules that best represent the targeted phenotype. Two of the biggest disadvantages of this approach are that: 1) the rules are rigid, meaning that they do not allow patients that have missing key data points to be included, and 2) these definitions are expert-driven and very time consuming/expensive to construct. Most recently, different ML-based approaches have gained traction, because they are data driven and allow more flexibility for patient inclusion [29]. Specifically, patients are assigned probability scores, rather than a binary label. In this work, we used a probabilistic score approach to examine racial bias in EHR data of disease phenotypes that impact older adult patients.

The National Institute on Aging has defined Alzheimer’s Disease and Related Dementias (ADRD) as a series of complex brain disorders that affect millions of Americans and as having a deleterious impact on individuals, their families, long-term care facilities, health care providers, and health care systems. The negative impact of ADRD on minority older adults cannot be overstated. In this study, we selected ADRD phenotypes as they impact all older adults, and especially in communities of color [30–33]. We utilize electronic phenotyping to characterize subgroup performance, which could lead to algorithmic biases (if any) in this context. A review of existing literature identified only one study by Straw and Wu [34]. that work Straw and Wu, present a sex-stratified analysis of machine learning models for liver disease prediction. In this work, instead of a sex-stratifed anlaysis, we build a more detailed and robust gender-stratified analysis to identify bias from a broader perspective, nicely providing an additional view of the problem than Straw and Wu. Additional prior research has examined racial bias in the context of dementia [35]. To our knowledge, this is the first study to evaluate the impact of racial subgroup performance, within probabilistic phenotyping models, on older adults in the context of mild cognitive impairment (MCI), Alzhimer’s disease, and Parkinson’s disease, Moreover, our study, which uses a larger EHR dataset, found very different conclusions on racial subgroup performance in dementia, thereby demonstrating the usefulness of our evaluation framework that was built for EHR data in the OMOP CDM format.

## OBJECTIVE

In this study, we characterized the racial subgroup in performance of probabilistic electronic phenotyping algorithms developed from EHR datasets. Without using race as a modeling variable, we hypothesized that (1) probabilistic algorithms perform differently for different racial groups, (2) the difference in performance is tied to data availability for different racial groups, and(3) not all algorithms show the same level of racial subgroup performance differences.

## MATERIALS AND METHODS

### Dataset

The dataset utilized for this work is from de-identified data from the Stanford Medicine Research data Repository (STARR), consisting of over three million patients with clinical data from 2008 until the end of 2018. This dataset has been converted to the OMOP CDM version 5.3 and used in multiple OHDSI studies over the years. Table 1 shows the overall demographics of the dataset. Of particular interest is the underlying racial distribution, which will be highlighted throughout the rest of this study.

**Table 1:**
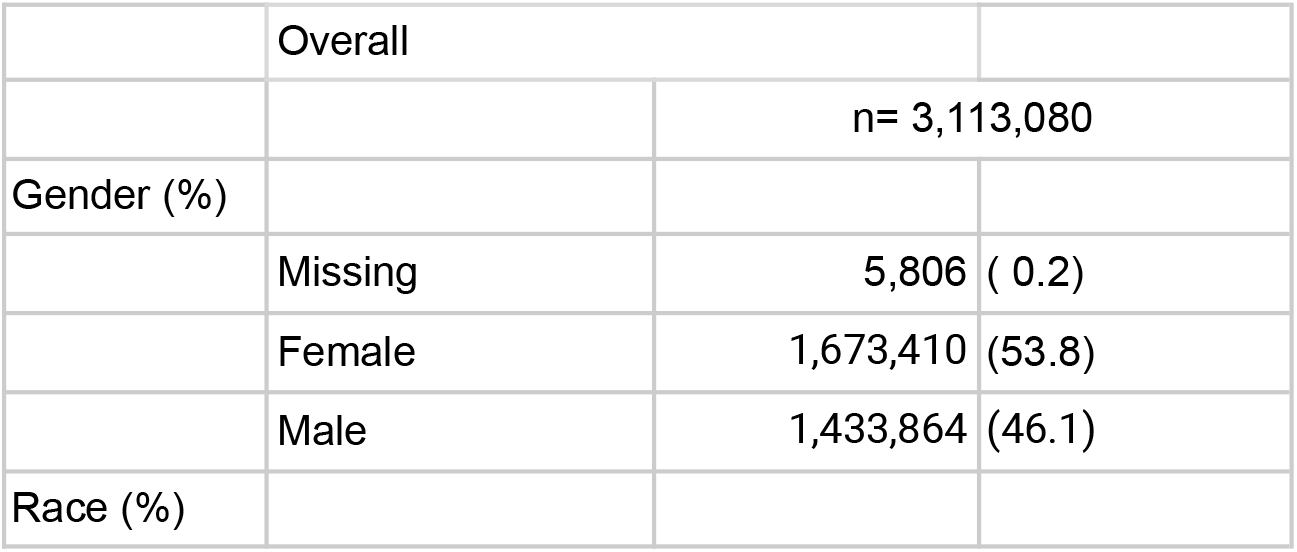
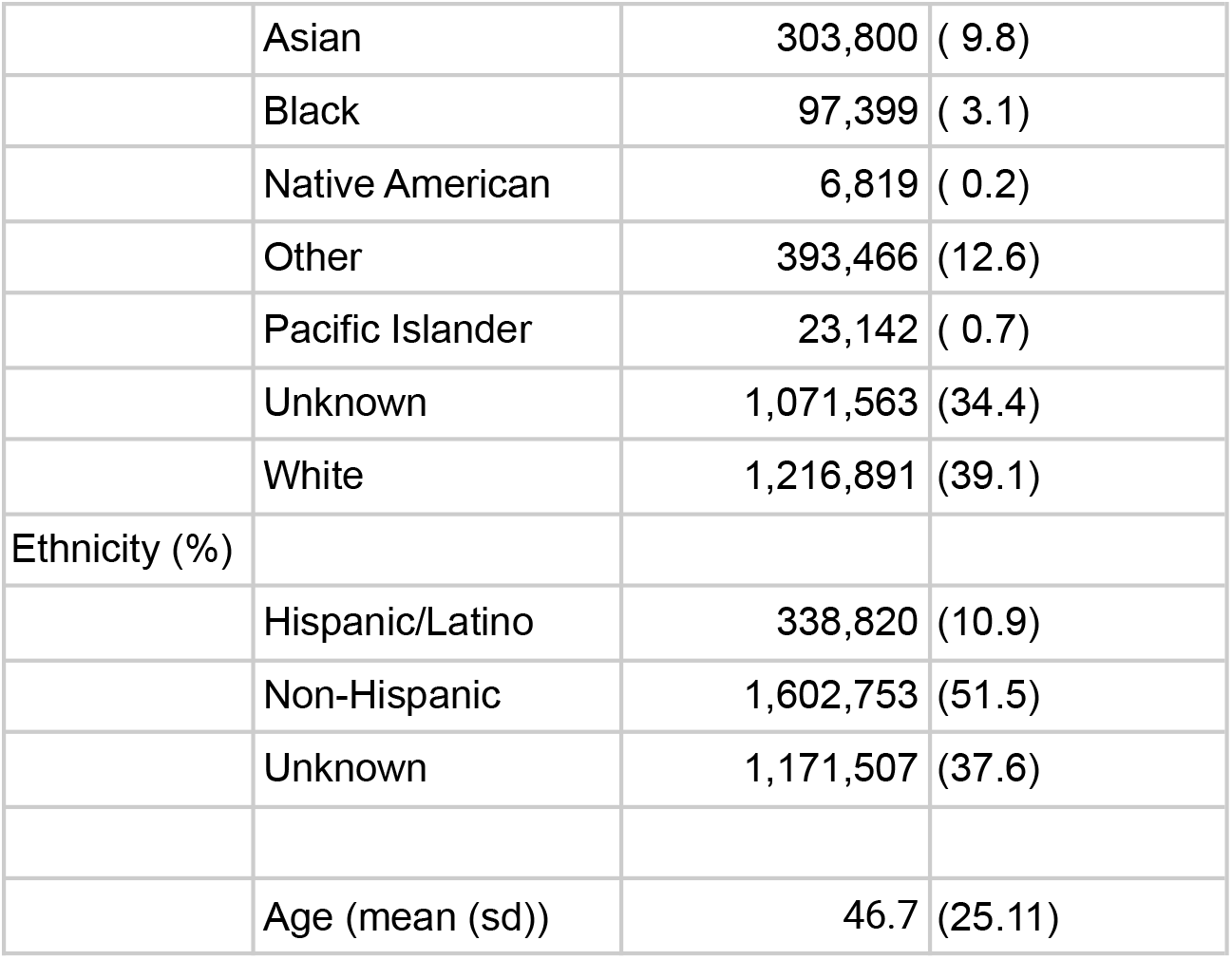
Demographics of patients from 2008-2018

|  |  |  |
| --- | --- | --- |
|  | Overall |  |
|  |  | n= 3,113,080 |
| Gender (%) |  |  |
|  | Missing | 5,806 ( 0.2) |
|  | Female | 1,673,410 (53.8) |
|  | Male | 1,433,864 (46.1) |
| Race (%) |  |  |

|  |  |  |  |
| --- | --- | --- | --- |
|  | Asian | 303,800 | ( 9.8) |
|  | Black | 97,399 | ( 3.1) |
|  | Native American | 6,819 | ( 0.2) |
|  | Other | 393,466 | (12.6) |
|  | Pacific Islander | 23,142 | ( 0.7) |
|  | Unknown | 1,071,563 | (34.4) |
|  | White | 1,216,891 | (39.1) |
| Ethnicity (%) |  |  |  |
|  | Hispanic/Latino | 338,820 | (10.9) |
|  | Non-Hispanic | 1,602,753 | (51.5) |
|  | Unknown | 1,171,507 | (37.6) |
|  | Age (mean (sd)) | 46.7 | (25.11) |

### Rule-based algorithms as our gold-standard

We used the rule-based phenotype algorithms validated and available on the HDR UK Phenotype library [36] for the following conditions: dementia [37] and Parkinson’s disease [38]. We adapted them into OHDSI ATLAS cohort definitions for their application on our OMOP-converted data. The rule-based phenotype definitions for mild cognitive impairment were adapted from Jongsiriyanyong and Limpawattana [39], and for Alzheimer’s disease we used the clinical definition by Holmes [40].

Manually curated and clinically validated sets of patients are more robust but also significantly more time and resource intensive. Thus community-approved rule-based definitions from organizations like PheKB [41] and CALIBER (now HDR UK Phenotype library) [42] have become the next best thing in order to computationally identify phenotypes. These rule-based definitions, while clinically validated, are quite rigid and show less flexibility than other approaches [28], this leads to many potential patients being excluded if certain codes or conditions are just not presented in their health record because of lack of coding or errors.

### Probabilistic phenotypes with APHRODITE

Instead of relying on rigid rules to define medical condition phenotypes, newer data driven approaches leverage machine learning to build statistical models to classify patients. These approaches have gained traction in the last few years [28], because they allow subjects to have a degree of probability of belonging to a phenotype, making them more flexible and able to catch people that have clinical codes missing or incomplete data. The methodology used for this work, APHRODITE, was designed with this flexibility in mind. Specifically, it relies on building statistical models for phenotypes based on an initial cohort of patients selected using high-precision keywords/clinical codes. The models were built using weak supervision where the patient’s entire clinical record up until the first appearance of the selected keyword/clinical code and. use weak supervision, where only an initial keyword/clinical code is needed and everything else is data driven. This approach was introduced by Agarwal et al. [43] and was made into an R package [44] which works on standardized data to the OMOP CDM format.

### Experimental framework

In order to identify racial subgroup performance variations within the probabilistic models, we built the following steps into a framework that we can later re-use for a wider variety of phenotypes. We started by selecting three of the most popular classical machine learning models to evaluate: LASSO [45], since regression-based classifiers are widely used for statistical learning purposes with EHR/medical data [46], Random Forest (RF) [47], and support vector machines (SVM) [48]. Note that the three models listed above are the ones supported by default by APHRODITE; however,any model supported by caret R package can also be included [49]. Next, we selected matched cases and controls to build our probabilistic models. The patients were matched by age, race, gender, and length of clinical record. We then stratified by the patient’s race for our multi-pronged evaluation, which consists of: traditional model (all races merged together), balanced model (we balance based on equal distribution of patients for each race), single race only model, and the leave-one-out combinations, which take one race out of the model building process in a systematic way. Following our usual practice, we used 75% of the data to train the model and a 25% unseen set to test the model, in addition to five-fold cross validation.

For evaluation, we used the traditional metrics: accuracy, which is the fraction of assignments the model identified correctly; sensitivity, which is the proportion of positives that are correctly identified; and specificity, which measures the proportion of negatives that are correctly identified. In addition, we used variation in order to measure difference between models in the following way:

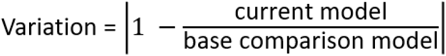

This measurement allowed us to evaluate the first three metrics in a similar context and show how different models are compared with each other. Note that while phenotyping algorithm performance is important, this is not the key point of this work, we present general model classification accuracy in order to put performance variance between phenotypes in context.

## RESULTS

### Phenotyping algorithms

First, we checked that the probabilistic phenotyping algorithms performed well when compared against their rule-based definitions. **Table 2** shows APHRODITE’s performance to select and identify the ‘gold-standard’ patients identified by the rule-based phenotype definitions.

**Table 2:** Rule-based and APHRODITE phenotype selection overlap.

| Phenotype | Cases identified by rule-based definition | Cases identified by APHRODITE keywords | Initial Overlap (%) | Classified by APHRODITE model (prob over 90%) | Total Overlap (%) |
| --- | --- | --- | --- | --- | --- |
| Dementia | 13,213 | 16,998 | 95.95% | 471 | 99.52% |
| Mild Cognitive Impairment | 7,915 | 8,292 | 94.14% | 399 | 99.18% |
| Alzheimer's disease | 11,401 | 12,828 | 89.04% | 1,137 | 99.02% |
| Parkinson's disease | 5,989 | 6,644 | 79.50% | 896 | 94.46% |

Our results show that APHRODITE and its probabilistic models are successful at identifying almost the same patients for each phenotype when compared to the rule-based definition, which served as our gold-standard.

**Table 3** shows the demographics of the patients identified for each phenotype. These results had a considerable impact on the resulting models and downstream evaluations.

**Table 3:** Patient demographics for the evaluated phenotypes.

| Phenotype | Cases | Controls | Gender |  | Race |  |  |  |  |  |
| --- | --- | --- | --- | --- | --- | --- | --- | --- | --- | --- |
|  |  |  | Female | Male | Asian | Black | Native Amer. | Pacific Island. | White | Unknown |
| Dementia | 16,998 | 16,998 | 56.22% | 43.78% | 11.07% | 4.96% | 0.27% | 0.80% | 60.33% | 22.57% |
| Mild Cognitive Impairment | 8,292 | 8,292 | 49.82% | 50.18% | 11.73% | 3.88% | 0.22% | 0.92% | 60.06% | 23.19% |
| Alzheimer's disease | 12,828 | 12,828 | 60.05% | 39.95% | 12.08% | 4.85% | 0.25% | 0.69% | 63.03% | 19.11% |
| Parkinson's disease | 6,644 | 6,644 | 39.30% | 60.70% | 12.06% | 1.34% | 0.23% | 0.32% | 63.11% | 22.95% |

A few things to note are that: 1) the racial distributions for some of the categories like Native American and Pacific Islanders are very low; 2) there were many patients listed as unknown race, which were removed from our evaluation.

Our evaluation framework produced over 1,200 plots and charts evaluating the performance of probabilistic models built under multiple conditions. **Figure 1** presents model classification accuracies for the four phenotypes and the three machine learning algorithms we utilized (i.e., 15 parameters) for the twelve different data subsets evaluated. This figure sets the precedent of the importance of the model variance evaluation and how it will change models from being potentially useful, to highly unreliable.

**Figure 1.**
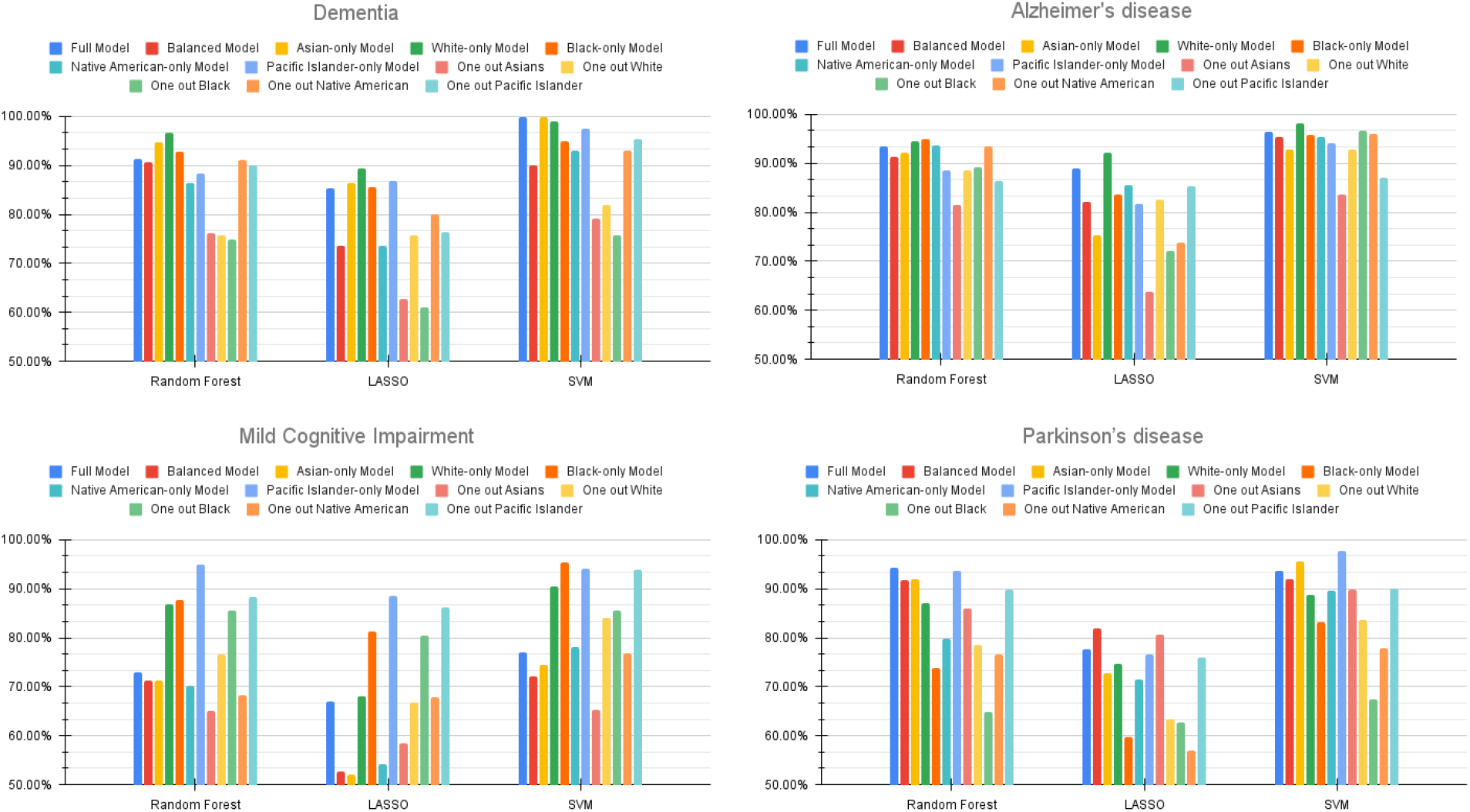
Model Classification Accuracy Result for all machine learning algorithms and all phenotypes.

These results demonstrate that most full models (i.e., the classical ones), which are the purple bars, have 70% to 90% classification accuracy for the given phenotype. These results are only for illustration purposes and to put the following figures of model variance in perspective. We are not trying to find the best performing models in general, but rather show their bias, when races are stratified.

### Evaluation scenario one: Building models with individual racial subgroups

In Figures 2 and 3 we show the classification and sensitivity variance for the random forest models between our models built for individual races and compared across all races evaluation. For example we built models using only White patients, and compared their performance when classifying patients from all other races. We used random forest as our choice algorithm to illustrate our results due to its solid average performance during our experimentation, and due to the more explainable nature of its models. Note that the same plots for specificity, as well as for the LASSO and SVM classifiers are provided as part of the supplemental appendix.

**Figure 2.**
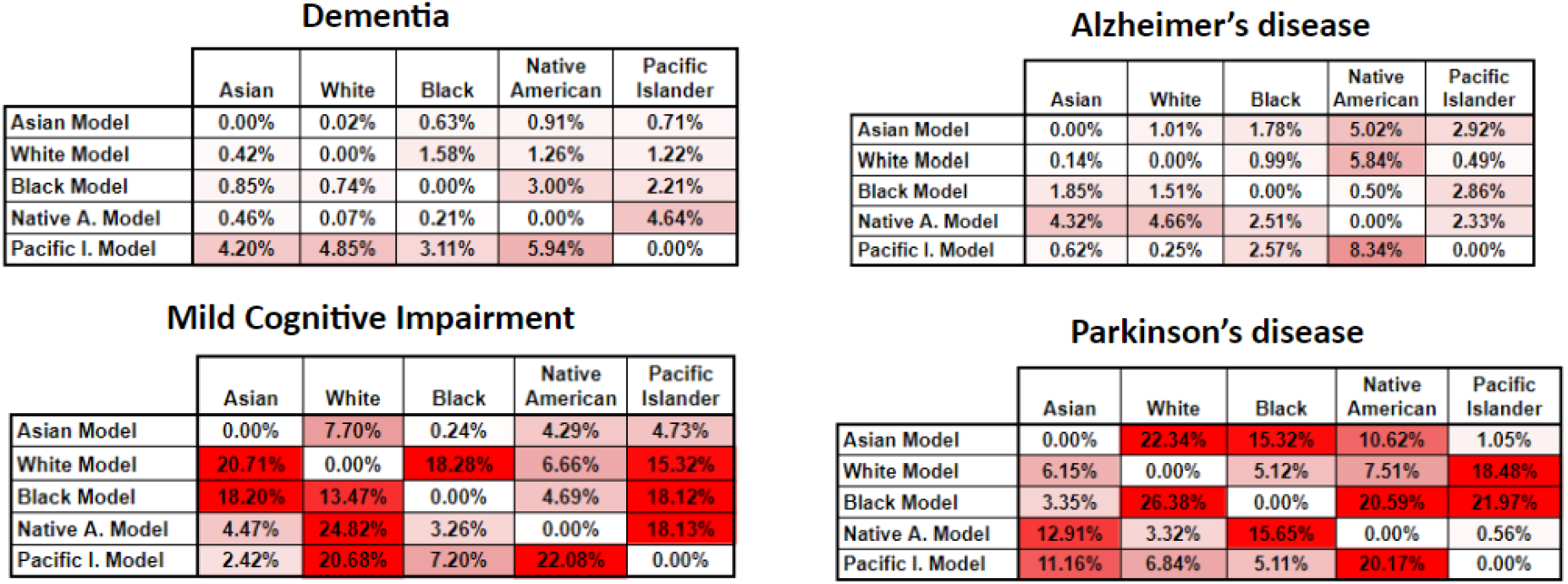
Classification accuracy variance for the Random Forest algorithm for our four phenotypes. In this heatmap, the higher tone of red a cell has, represents bigger variance from the baseline, or in other words more bias.

**Figure 3.**
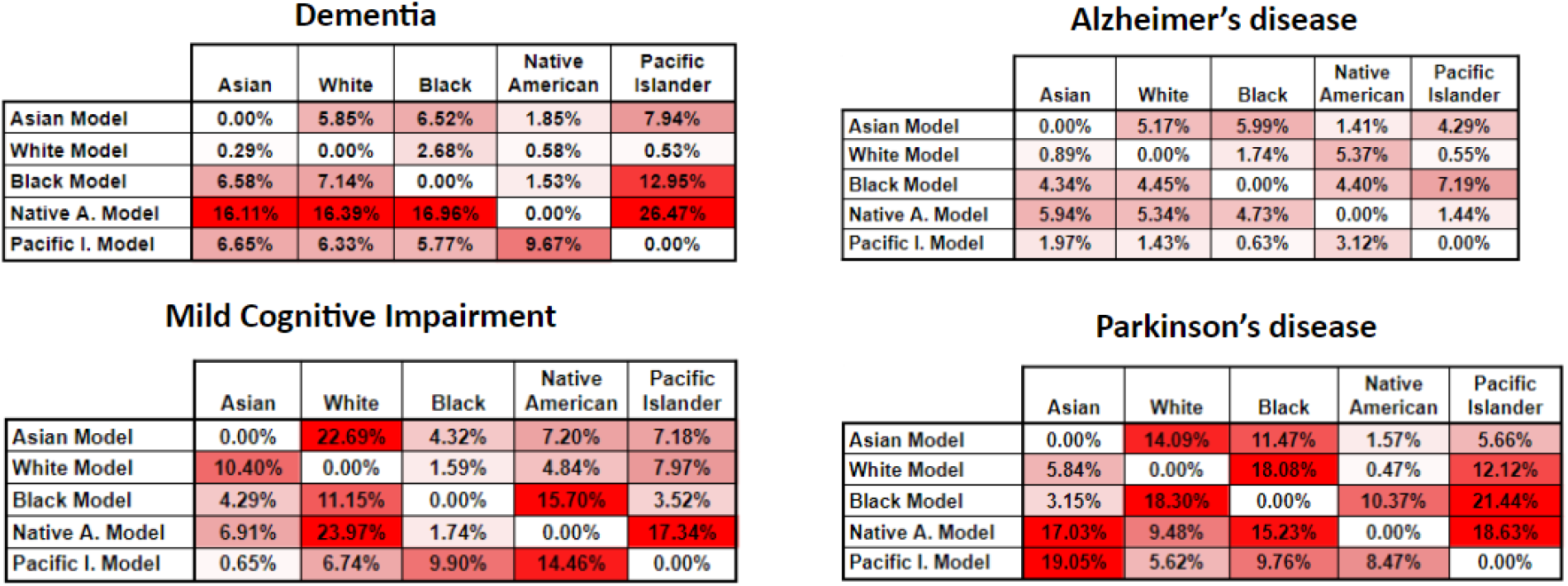
Classification sensitivity variance for the random forest algorithm for our four phenotypes. In this heatmap, the higher tone of red a cell has, represents bigger variance from the baseline, or in other words more bias.

Figure 2 shows some very interesting results for the Dementia and Alzheimer’s disease phenotypes, illustrating that the variance between models is not that pronounced (less than 8% at the worst case). This shows that models built with the individual races are generalizable enough across races, at least for these two phenotypes. For the remaining three phenotypes the variance increases up to 27%, rendering a classification model with an accuracy of less than 80%, almost equal to random picking. These results showcase the need to evaluate these combinations carefully, particularly before deployment of any phenotyping model.

The sensitivity variance for the models on Figure 3 shows a similar trend as Figure 2–i.e., less variance for the Dementia and Alzheimer’s phenotypes. However, there is a considerable increase in the variance of the Native American model, most likely due to the fact that this racial group is highly underrepresented in the dataset used; thus this model is unable to properly generalize across races, particularly when measuring sensitivity.

### Evaluation scenario two: Full models, balanced models, and leave one-out

We now switch to evaluate the models in a more traditional sense of using a model with all data available–one that balances the classes but limits the number of samples to the minimum available in any given class. Note that we did not use SMOTE [50] or any sampling techniques in this work, as this is not ideal when using clinical data [51,52], since depending on the method used, it adds non-representative extra data. The other scenarios we evaluated include leaving one class out in the model building process. We then applied the built models to the individual classes of patients in the testing set (fully unseen patients). Figures 4 and 5 report these results for classification accuracy and sensitivity. Additional figures for the other machine learning learning models and metrics are available in the supplemental appendix.

**Figure 4.**
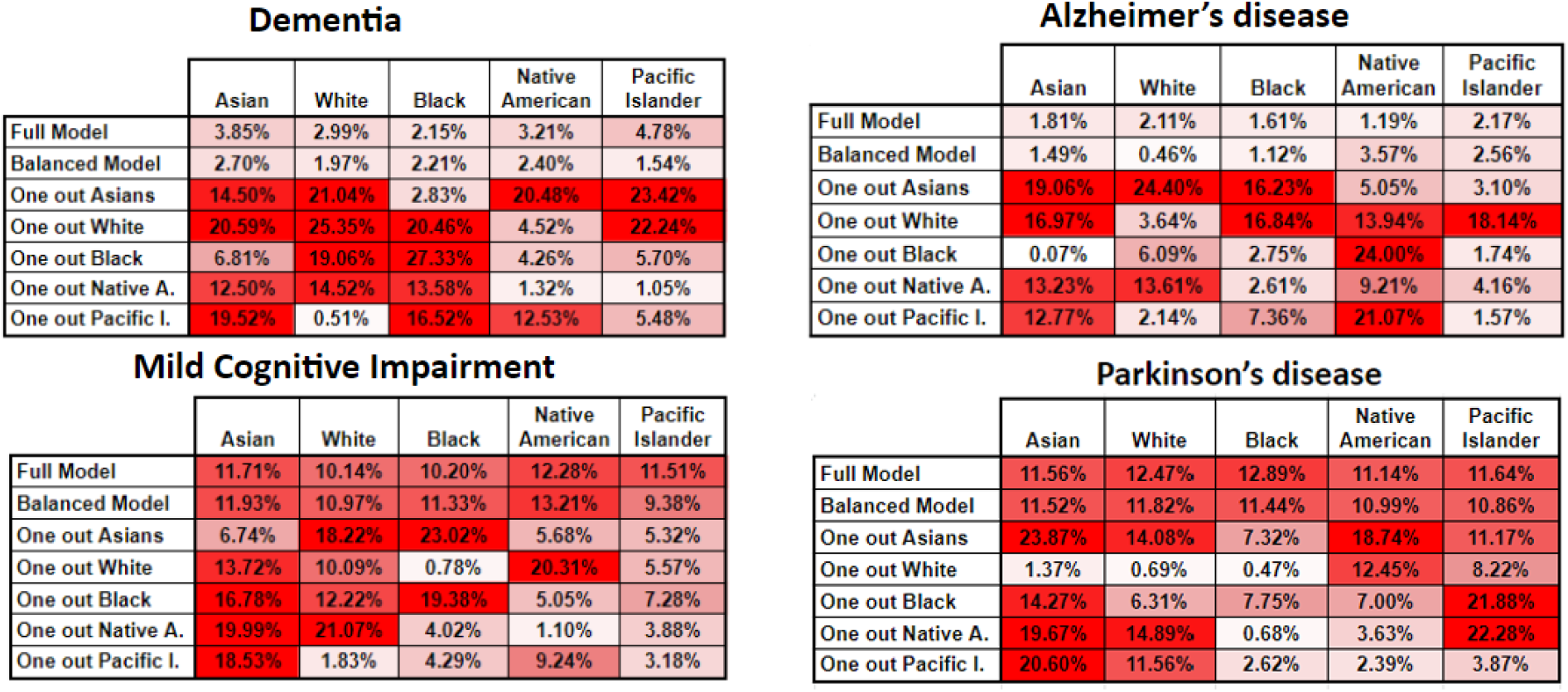
Classification accuracy variance for the Random Forest algorithm for our four phenotypes. In this heatmap, the higher tone of red a cell has, represents bigger variance from the baseline, or in other words more bias.

**Figure 5.**
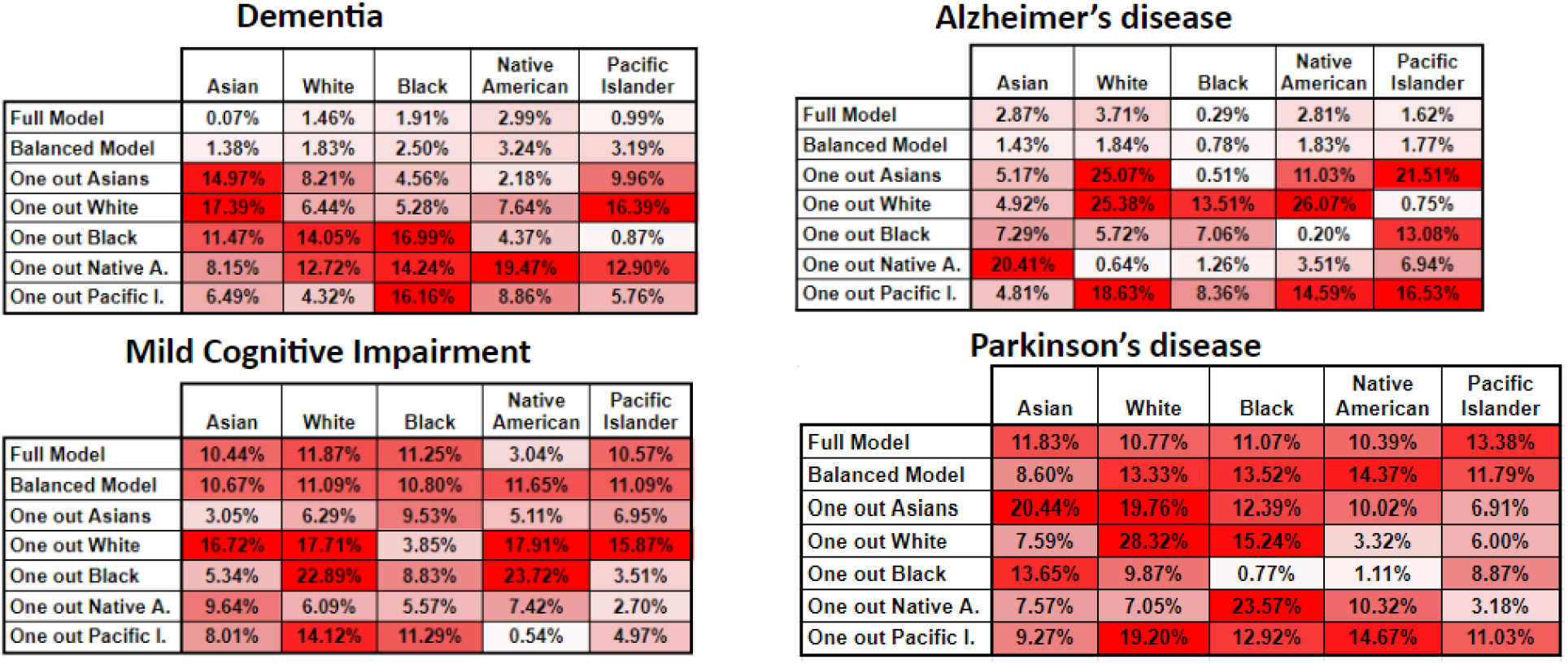
Classification sensitivity variance for the Random Forest algorithm for our four phenotypes. In this heatmap, the higher tone of red a cell has, represents bigger variance from the baseline, or in other words more bias.

We again see very similar patterns for the Dementia and Alzheimer’s disease phenotypes, where the variance is quite low. One thing to note is that there is always variance here as the calculation is performed on the baseline model performance only classifying the given class (on the unseen test set) versus the baseline model performance of all classes on the unseen test set. One very interesting result here is that for the phenotype models with less subgroup variance, taking out certain classes brings their overall performance down by considerable amounts (up to 26%) sometimes, which could be mostly due to removing the data-dominant race. In other scenarios the accuracy variance is not that high, particularly when classifying the races with very small representation in the original models.

Figure 5 shows that classification sensitivity variance is also affected in a similar way as for the classification accuracy in almost the exact same way with the variance trends being very similar. This also reinforces the original finding, namely that two of the phenotypes show small variance, across our experimental evaluation and the other three have different, but increasing, degrees of variance. These results demonstrate that probabilistic phenotype models need to be carefully examined and improved before they can be used in a clinical setting.

## DISCUSSION

We designed our experimental framework to provide a fully-automated and standardized (on top of the OMOP CDM and using APHRODITE R package) way to demonstrate if any given probabilistic phenotyping model has racial subgroup variance and estimate how much. As shown in the results sections, we have two different evaluation scenarios, which build probabilistic models in different stratified ways to provide flexibility and insight into how the differently built models will vary given popular machine learning metrics. Note that any identified anomaly in subgroup performance does not automatically translate into the algorithm leading ‘harming’ subgroups, as addressing or ‘fixing’ those anomalies might actually produce worse performing algorithms for all subgroups as found by Pfohl et al. in [53]. In some cases having different algorithms for subgroups or a human in the loop might [54] be a better approach to not affect any group of patients.

For the phenotypes of our case study, the first scenario clearly demonstrates that two phenotypes: dementia and Alzheimer’s disease present the least amount of sub group variance, (Figures 2 and 3), both in terms of classification accuracy and sensitivity. This is highlighted by their two variance figures showing the least amount of red cells. These figures are designed as heatmaps to visually highlight the severity of the variance and place the attention of the researcher on the more relevant sections. Racial representation in those cohorts could be one possible reason these phenotypes show less subgroup variance, but while they have some of the highest numbers of cases used for training (Table 2), their racial representation is nearly the same as in the whole dataset. An interesting observation is that accuracy shows very little variance whereas the sensitivity results show a higher amount of it. This might indicate that while the stratified models do well as a whole, there might be additional small variance when trying to detect the positive class. However, this can also be explained as an artifact of building the model with the Native American patients, which had the least representation in the full dataset (> 0.25%). A common solution to address underrepresentation has been to oversample this class, or undersample some of the others. However, our second scenario shows that this does not significantly improve the level of subgroup variance, as other researchers have shown, at least for predictive tasks [51,52]. Rather, we recommend using federated learning, as this is starting to be more accepted in large research networks [55], or use ensembles using multiple datasets, when available within a single site or research facility [56]. The second evaluation scenario shows that the full model and the balanced model perform very consistently for the phenotypes of dementia and Alzheimer’s disease, which have less subgroup variance. However, it also shows very striking results on the leave-one-out models, particularly when removing the racial groups with the larger representations, and when removing a particular racial group and then trying to classify only patients of that group (Figures 4 and 5). These results show the need to consider such detailed analyses before trying to use any of these models in clinical practice.

Regarding the phenotypes—mild cognitive impairment, and Parkinson’s disease— with more subgroup variance, we observed some very dramatic variance changes (average of 10%) between most of the experimental models. This indicates that those phenotypes are quite sensitive to any of the racial groups being removed, particularly shown by the leave-one-out models from the second scenario (Figures 4 and 5). These figures also show that even for the full-model and balanced scenarios, predicting on individual classes brings considerable accuracy and sensitivity variance. These findings translate to how sensitive these models are to any type of shift in the underlying dataset that is used to train the model and how to evaluate them. These results strongly demonstrate the need for rigorous experimental evaluation before any kind of deployment or testing in production environments (e.g. hospitals). While there are plenty of experimental evaluations to analyze, our framework automates the work for researchers, and it only needs human interpretation of its findings.

The limitations of this work are the following: we evaluated three machine learning algorithms based on their popularity and level of use within the field. However, with new algorithms constantly being introduced, the results could vary dramatically when other algorithms are introduced. We decided to keep the same algorithms as in our previous works [44,57] since we know how those perform to build probabilistic models using APHRODITE. The flexibility behind APHRODITE, allows for other models to be evaluated within the presented framework as long as they have an R package available. We decided to only evaluate the variance metric as it gives a stronger and more interpretable signal on how the model differs from each other. For this study we needed to keep the number of evaluations and experiments to a reasonable amount to be able to explain this work and its merits. However, any other metric can be configured into our framework. Lastly, our case study phenotypes were evaluated on a single dataset, and, in future work, we plan to fully leverage the OHDSI community to conduct a network study examining racial bias of phenotypic algorithms [58,59]. One major item to note is that self-reported race is usually error prone and very often incomplete (missing in up to 23% of the patients selected for the phenotypes evaluated), these factors could lead to some of the results being artifacts of this phenomenon.

## CONCLUSION

As we have demonstrated in this work, subgroup performance variance can certainly be found in probabilistic phenotype algorithms that categorize older adults, particularly for phenotypes like mild cognitive impairment, and Parkinson’s disease. As a result of this subgroup variance, models perform up to 30% worse under our multiple model building scenarios. Thus it is critical for institutions to extensively test and rigorously evaluate their phenotyping models. We found that some phenotypes like dementia and Alzheimer’s disease were more resistant to this subgroup variance as indicated by their very small variance under all of our testing scenarios, meaning that these models could be potentially used safely. Rigorous testing allows researchers to be more confident of the performance of these models under different racial distributions. Our main contribution is the framework to fully automate this process when the institution has data in the OMOP CDM and can run our extension to the APHRODITE package. This work is particularly important as biomedical scientists and medical professionals strive to make informed conclusions and diagnoses of older adult patients.

## Data Availability

All data produced in the present study are available upon reasonable request to the authors

## AUTHOR CONTRIBUTIONS

Conception and design: All authors. Data analysis: JMB. Data interpretation: JMB. Drafting the manuscript: All authors. Revising the manuscript: All authors. Approval of submitted version: All authors. Accountability of own contributions: All authors.

## DATA AVAILABILITY

The EHR data used in this study cannot be shared for ethical/privacy reasons. All code is available in the following location: https://github.com/OHDSI/Aphrodite

## ACKNOWLEDGEMENTS

The authors would like to thank Jessica Moon, PhD, PMP for her comments and proofreading of this manuscript.

